# Effectiveness of the Smoke Free App for Smoking Cessation – Results of RAUCHFREI, a Randomised, Double-blind, Controlled, Two-arm, Parallel, Nationwide, Decentralised, Fully Remote Clinical Trial in Germany

**DOI:** 10.64898/2026.03.17.26348617

**Authors:** Lucas Keller, Anne Schraplau, Patrick Timpel, Tonio Schönfelder, Sandy Scheibe, Ria Heinrich, Jonathan B Bricker, Jamie Brown, Felix Naughton, Tobias Raupach, Robert West, Benedikt Pontes da Silva, Caroline Schmidt-Lucke, David Crane

## Abstract

**Objectives:** Uptake of evidence-based smoking cessation support remains limited. Digital interventions offer the prospect of scalable and highly accessible support. Smoke Free, a digital mobile application using established behaviour change techniques, has shown promise, but no large-scale randomised controlled efficacy trial has yet been conducted. We assessed its effectiveness for smoking cessation.

**Design:** In this prospective, randomised, controlled, two-arm, parallel clinical trial with 6-month follow-up, study personnel and patients were blinded.

**Setting:** The trial was conducted nationwide in Germany, utilising a decentralised, fully remote trial design. Enrolment took place digitally after receiving brief advice from a healthcare professional, following guidelines for primary care.

**Participants:** Out of a volunteer sample of 1850 patients assessed for eligibility, 1466 adult cigarette smokers who had at least moderate cigarette dependence (F17.2, FTCD≥3) were recruited between August 2023 and February 2024; 84.1% (1233 participants) completed the primary outcome measure.

**Interventions:** The intervention group (IG) received the Smoke Free app including behaviour-change missions and gamification elements, while the control group (CG) received a text-only cessation information app. Both groups received brief advice from a healthcare professional.

**Main outcome measures:** The prespecified primary outcome was self-reported 7-day point-prevalence abstinence from combustible tobacco at 6 months post-randomisation; secondary outcomes included biochemical validation of abstinence in participants providing a saliva sample (59% of eligible participants).

**Results:** Self-reported abstinence (primary outcome) was significantly higher in the IG compared with the CG (283 [39.3%] vs. 182 [24.4%], OR=2.01, 95% CI 1.60 to 2.50, p<0.0001). The NNT was 6.7 (5.1 to 9.8). The effect was consistent with biochemical validation (OR=1.76, 95% CI 1.27 to 2.44, p<0.0001) and across secondary outcomes and sensitivity analyses. The 6-month follow-up rates for the primary outcome did not differ between groups (IG: 601 [83.5%]; CG: 632 [84.7%]; p=0.52). Eighty-four serious adverse events were reported by 75 participants (IG: 31, 4.3%; CG: 44, 5.9%; p=0.53); none were treatment-related.

**Conclusions:** The Smoke Free app is effective for aiding smoking cessation in at least moderately dependent cigarette smokers compared with an informational app when provided as an adjunct to brief advice from a healthcare professional.

**Trial registration:** The trial was registered with the German Clinical Trials Register (DRKS00031140).

**Funding:** Smoke Free 23 GmbH (for-profit company).

---

Many smokers struggle to quit, and use of evidence-based cessation assistance remains limited. The International Tobacco Control Policy Evaluation Project, a longitudinal multi-country effort evaluating tobacco control policies, has documented substantial cross-national variation but persistent gaps in assistance uptake. In a survey of smokers of eight European countries, the majority (56–84%) in every country except in England undertook their most recent quit attempt unaided, with low use of pharmacotherapy, quitlines, or other services.(1) In Germany, only a small minority report the use of at least one evidence-based support method during their last attempt(2) and 12-month quit attempts have declined by more than two-thirds over the past seven years.(3)

Mobile digital applications (smartphone apps) are routinely used by billions of people worldwide and numerous smoking cessation apps have been developed.(4,5) However, there is limited evidence on the effectiveness of specific apps and they vary considerably in terms of the behaviour change techniques that they use or the way that they deliver these.(6) Evaluations are mostly hampered by low follow-up rates,(4) with a notable exception.(7) Overall, there is uncertainty about the effectiveness of app interventions without concurrent pharmacotherapy,(4) to date, only one app offered without pharmacotherapy has proven efficacious in a larger clinical investigation.(7,8) This gap in knowledge is particularly significant given the immense potential for apps to become scalable behavioural interventions, reaching many smokers cost-effectively, especially those who may be unable or unwilling to physically attend human-delivered services regularly.(5) This becomes especially important when scaling theory- and evidence-based solutions to the estimated 1.3 billion smokers worldwide.(9)

The present study was designed to determine whether the Smoke Free smartphone app, a theory- and evidence-based intervention, could significantly improve quit rates when used as an adjunct to standard clinical care. The intervention was delivered within the Digital Health Application (DiGA) fast-track system in the German healthcare system. DiGAs are digital interventions vetted by competent authorities that can be prescribed to patients by doctors and psychotherapists while costs are covered by health insurance. DiGAs must show their clinical effectiveness in a randomised controlled trial (RCT) as well as fulfil requirements regarding interoperability, data security, and data protection.(10)

Recent studies on the Smoke Free app have provided first promising results regarding its effectiveness for smoking cessation. One RCT showed that merely offering the app to smokers motivated to quit did not significantly improve cessation rates. However, restricting the analyses to participants who downloaded the app revealed a higher self-reported 6-month continuous abstinence compared to those not offered the app (12.7% vs. 7.0%, p<0.001).(11) Two earlier RCTs with more than 70,000 participants combined showed that the full version of the Smoke Free app significantly increased self-reported continuous abstinence compared to a reduced version but these trials had very low follow-up rates (<11%).(12,13) Another study showed increased user engagement through the introduction of gamification elements.(14) A narrative description and a full overview of the features and behaviour change techniques used by the Smoke Free app can be found in Appendix A.

This RCT evaluated the effectiveness of the Smoke Free smartphone app within the German healthcare system in promoting self-reported 6⍰month abstinence among at least moderately dependent smokers, compared with an informational app, when both were used alongside brief professional advice.

## Methods

### Study design

This was a double-blind, controlled, two-arm, parallel, nationwide, decentralised, fully remote clinical trial with stratified block randomisation and prespecified primary and secondary endpoints. Participants from all 16 states of Germany formed a community sample that was recruited over internet search ads, within app stores, and with flyers and print ads. Ethics approval (#Eth-61/22) was obtained from the ethics committee of the Medical Association Berlin (Germany) and participants were enrolled into the study after confirming their identity and giving written informed consent. The trial was registered with the German Clinical Trials Register (https://drks.de/search/en/trial/DRKS00031140) on Feb 02, 2023, before the inclusion of the first participant. The study was run by MEDIACC GmbH, a clinical research organisation under the supervision of CSL.

### Participants

Participants were adult smokers with a tobacco dependency (ICD-10 F17.2). This was assessed through online screening (using the Fagerström Test for Cigarette Dependence, FTCD(15)). Table 1 gives an overview of the eligibility criteria which were confirmed by medically trained study personnel during a video consultation.

**Table 1:** Eligibility criteria.

| Inclusion criteria | Exclusion criteria |
| --- | --- |
| ☐ (at least) moderate tobacco dependency (FTCD≥3)<br>☐ Motivated to quit (currently)<br>☐ Compatible smartphone (Android≥7.0; iOS≥14.0)<br>☐ Willingness not to use another stop smoking app during the trial period<br>☐ Residency in Germany and valid contact data<br>☐ Sufficient language skills (B2 German) | ☐ Below the minimum (18 years) or above the maximum (65 years) age<br>☐ Specific comorbidities<br>○ Diagnosed substance use disorders (untreated or currently in treatment)<br>○ Acute and severe psychiatric disorders or suicidality<br>○ Cognitive impairments or dementia<br>○ Impending hospitalisation during the trial<br>☐ Living together with someone using Smoke Free or also participating<br>☐ Having used Smoke Free in the past 12 months |

Sex (male or female) was self-reported. Written informed consent was obtained after confirmation of eligibility and the ABC brief intervention via an electronic signature of the informed consent form.

### Randomisation and masking

Randomisation was computer-generated using block sizes of eight, stratified by (1) tobacco dependence (medium-high, FTCD≤6, vs. very high, FTCD≥7) and SES (income and education composite score: low, ≤3.5 out of 11, vs. medium-high, ≥4 out of 11; see Table A3 in Appendix B). The concealed allocation sequence created four stratification cells and was accessible only to an IT service provider that designed the algorithm and the trial monitor. After entering a study identification code, participants were randomised automatically within the app at a time and place of their choosing, without involvement of study personnel.

### Procedures

All eligible participants received the ABC brief intervention (ask, brief advice, cessation support (16,17)) provided by the medically trained staff at the end of the screening video consultation. This took about 3–5 minutes and confirmed the participants’ motivation to quit. Although the ABC intervention mentioned the use of medication or other forms of assistance, participants were not offered medication or other forms of assistance as part of the trial. Participants then had seven days to sign the consent form, after which they received the baseline questionnaire on the following morning. The baseline questionnaire comprised questions regarding demographics (eg, level of education, sex), smoking-related questions (eg, past quit attempts, confidence to quit, smoking status), and exploratory outcomes not pertinent to the current report. At the end of the baseline questionnaire, participants were instructed how to install the app and to enter their code.

Participants in the intervention group (IG) received access to the full app for 6 months. Key features of the full app include interactive behaviour change tools, progress tracking, and personalised motivational support (a summary and an assessment of included behaviour change techniques can be found in Appendix A). Participants in the control group (CG) received access to a reduced, minimal support version of the app for 6 months that was identical in design on a smartphone’s home screen. The reduced version offered static text-based smoking cessation information modelled on brochures/flyers common in GP offices but lacked any interactive content (see Appendix A).

After 30, 90, and 180 days, participants received invitations via email and text messages to fill out follow-up (FU) questionnaires: FU1, FU2, and FU3, respectively. The online questionnaires were available for 14 days each. Participants received reminders after 1, 3, and 5 days, and were called by members of the study team if they had not completed the questionnaire after a week. Participants received €20 each for completing the first two follow-ups. For completing the last follow-up, participants received €30 and an extra €10 if they completed it within the first 24 hours. Participants could choose whether to receive this as a voucher or to donate to one of four charities. Moreover, every at this point remaining participant received a noncontingent reward (a €5 note) 2 to 3 days before FU3, together with a postcard informing them of the upcoming measure. These approaches have been shown to increase adherence and minimise loss to follow-up in previous studies.(18) Table A4 in Appendix C gives an overview over the assessments at each visit.

### Outcomes

The primary outcome, self-reported 7-day point-prevalence abstinence (PPA) from combustible tobacco products (ie, smoking abstinence), was assessed as the first question of each online questionnaire (baseline and every follow-up). A missing value was counted as a return to smoking (ie, missing-equals-smoking imputation). In line with recommendations,(19) it was followed by questions assessing the 7-day PPA from electronic nicotine delivery systems (ENDS), heated tobacco products, and non-combustible tobacco products. This allows the reporting of abstinence from combustible tobacco products (smoking abstinence; primary outcome), abstinence from combustible and smokeless tobacco products (ie, tobacco abstinence; exploratory outcome), and abstinence from all tobacco products and ENDS (ie, nicotine abstinence; exploratory outcome).

The first secondary outcome, biochemically validated abstinence, was measured with a cotinine saliva test (cut-off: 20 ng/mL) at FU3. Participants reporting nicotine abstinence were invited to take part. Saliva samples were collected under supervision in a video consultation after confirming the participant’s identity to ensure sample integrity (more detail in Appendix D). Participants with inconclusive or missing cotinine results were considered smokers. To count as abstinent in the second secondary outcome, prolonged abstinence, participants had to complete and report smoking abstinence at each follow-up (ie, repeated PPA). If they had missed or reported having smoked in the last 7 days in one of the assessments, they were classified as having returned to smoking. The last secondary outcome, continuous smoking abstinence, was operationalised without a grace period but with slips, meaning that participants must have had no more than 5 cigarettes since their target quit day to count as abstinent.(20) This was assessed at every follow-up.

Usage data of the app were collected from participants in the IG. We measured app openings and the number of quit attempts recorded in the app. Moreover, participants of both groups were asked whether they would recommend the app using the net promoter score (NPS).(21) It is measured on a scale from 0 to 10, classifying responses of 9 and 10 as promoters. Lastly, participants of both groups were asked how satisfied they were with the app on a scale from 1 (not at all) to 5 (very).

Safety and adverse events were assessed at every follow-up questionnaire with the study team contacting participants in case further information was needed to assess the severity of the event and possible relation to the intervention. In case a participant whose responses to the questionnaire suggested at least one serious adverse event could not be reached after at least four contact attempts via at least two communication channels, the adverse event was automatically classified as serious.

### Power calculation

The sample size was calculated assuming ⍰=0.05 (two-tailed), 1−β=0.80, and a 6% intervention difference based on conservative assessments of the impact of digital interventions and brief advice, respectively (18% vs. 12%; see Appendix E for more detail). At the request of the German authority regulating DiGAs, the resulting number was further increased to counter an assumed attrition rate of 34%. This led to an adjusted target sample size of 1,442 participants.

### Statistical Analysis

All analyses of the prespecified primary and secondary endpoints followed a statistical analysis plan finalised before data collection began. Additional exploratory analyses were conducted post-hoc as described below. Efficacy analyses followed the intention-to-treat (ITT) principle, comprising every randomised participant in the group to which they were allocated. For all abstinence outcomes, participants with missing smoking status were, in accordance with the Russell Standard,(20) classified as smokers. Primary, secondary, and exploratory outcomes were analysed using logistic regression, producing odds ratios (ORs) with 95% confidence intervals. A second model, exploratorily, adjusted for the socioeconomic stratum, sex, age, age at smoking initiation, cigarettes smoked per day, desire to quit (0–10 scale), confidence to quit (0– 10 scale), and strength of tobacco dependence (continuous FTCD-score); all assessed at baseline.

Exploratory post-hoc analyses examined the descriptive depiction of associations between app engagement metrics and outcomes. Similarly, drop-out or different steps in the cotinine test funnel were compared between groups using Fisher’s exact test. Satisfaction with the app and NPS-scores were compared using t-tests. Safety outcomes were summarised descriptively; treatment-relatedness was adjudicated blind to allocation.

Furthermore, explorative sensitivity analyses for the primary endpoint were conducted. First, the analysis was repeated using multiple imputation with predictive mean matching. For this, 200 imputations with 10 iterations while controlling for all relevant baseline characteristics and intermediary abstinence reports were used and then fed into a logistic regression model testing the intervention effect adjusted for the baseline variables. Second, both treatment groups were compared using Fisher’s exact tests within each level of the stratification variables (tobacco dependence, SES) and sex, leading to six comparisons. Third, Fisher’s exact tests should have been conducted for groups of pharmacotherapy users, but the number of users was too low to conduct these analyses. Fourth, Fisher’s exact tests were conducted assuming all active withdrawals were abstinent or only active withdrawals in the CG were abstinent instead of the missing-equals-smoking imputation.

All tests were two-sided (α=0.05) with no multiplicity adjustment for secondary/exploratory outcomes. Analyses used JASP 0.19.3.(22) The Paper Authoring Tool,(23) an online tool that secures the inclusion of all essential methodological and outcome details in a structured, machine-readable format, was used in preparation of this manuscript.

### Patient and public involvement

Patients contributed to the development and refinement of the Smoke Free app through formative user research and usability testing conducted prior to study initiation.(24) Feedback from current and former smokers informed content design and feature prioritisation to maximise relevance, engagement, and usability. No patients were directly involved in the design, recruitment, or conduct of the study.

## Results

Between Aug 04, 2023, and Feb 19, 2024, 1,850 participants took part in online video consultations with the study team to assess their eligibility. Of those, 233 (12.6%) were classified as not eligible by the study team, 88 (4.8%) did not provide their informed consent, and 63 (3.4%) did not continue to activate the app and start the randomisation process after signing the informed consent. This means that a total of 1,466 participants were randomly assigned to the IG (n=720) or CG (n=746), see Figure 1. After randomisation, no participant was excluded but some participants did not complete the follow-up questionnaires or actively withdrew their consent for further follow-ups. Both groups were retained for analyses assuming a return to smoking. The number of active withdrawals was different between groups (OR=0.56, 95% CI 0.34–0.92, p=0.0195), with fewer participants in the IG (28, 3.9%) than the CG (50, 6.7%). Total loss to follow-up was similar between IG and CG (16.5% vs. 15.3%, p=0.52). A total of 1,233 participants (84.1%) completed at least the primary outcome in the final follow-up questionnaire after 6 months.

**Figure 1.**
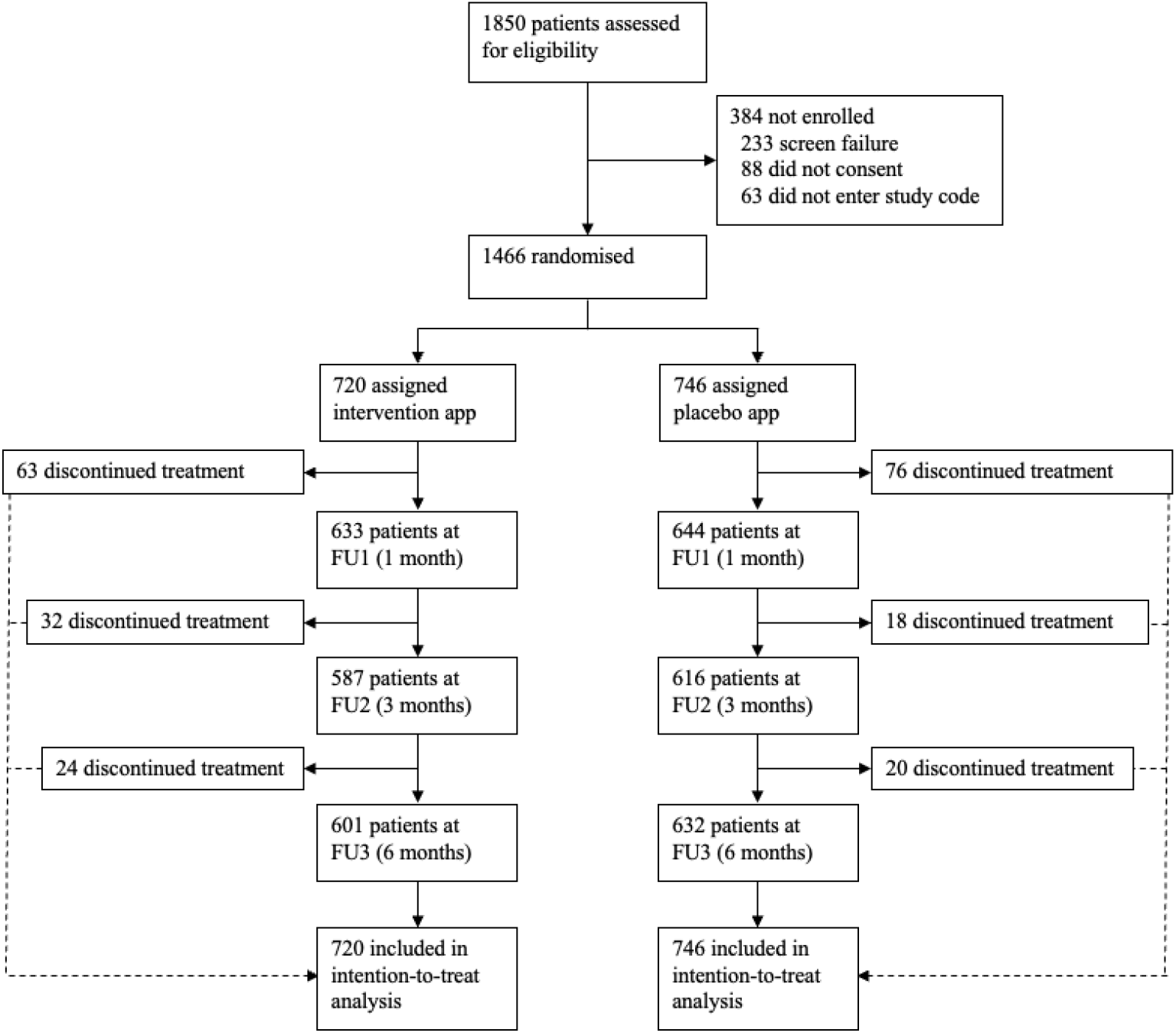
Trial profile.

Baseline characteristics of participants are presented in Table 2. The median age of participants was 37 (30–45) years and there were more female (887 [60.5%]) than male participants (579 [39.5%]). The baseline characteristics were similar between both groups.

**Table 2:** Baseline characteristics of the ITT population.

|  | IG (n=720) | CG (n=746) |
| --- | --- | --- |
| Sex |  |  |
| Female | 433 (60.1%) | 454 (60.9%) |
| Male | 287 (39.9%) | 292 (39.1%) |
| Age (years) | 37 (31–44) | 37 (29–45) |
| In a relationship | 472 (65.6%) | 492 (66.0%) |
| Education |  |  |
| no school-leaving certificate | 2 (0.3%) | 7 (0.9%) |
| basic secondary school leaving certificate | 48 (6.7%) | 34 (4.6%) |
| intermediate secondary school leaving certificate | 209 (29.0%) | 218 (29.2%) |
| universities of applied sciences entrance qualification | 141 (19.6%) | 143 (19.2%) |
| general university entrance qualification | 320 (44.4%) | 344 (46.1%) |
| SES-stratum (composite score of education and income) |  |  |
| low | 66 (9.2%) | 56 (7.5%) |
| high | 654 (90.8%) | 690 (92.5%) |
| Smoking-related variables |  |  |
| Age at which smoking was initiated (years) | 16 (14–22) | 16 (14–23) |
| Cigarettes smoked per day | 18.3 (6.8) | 18.2 (7.0) |
| Living in a household with other smokers | 235 (32.6%) | 231 (31.0%) |
| Previous quit attempts | 4.5 (6.1) | 4.4 (5.5) |
| Experience of withdrawal symptoms in prev. quit attempts | 624 (86.7%) | 654 (87.7%) |
| Past usage of behavioural support | 123 (17.1%) | 102 (13.7%) |
| Past usage of nicotine replacement therapy | 42 (5.8%) | 45 (6.0%) |
| Past usage of Bupropion | 10 (1.4%) | 11 (1.5%) |
| Past usage of Varenicline | 11 (1.5%) | 7 (0.9%) |
| Past usage of a stop-smoking app | 61 (8.5%) | 62 (8.3%) |
| Desire to quit (0–10) | 8.4 (1.5) | 8.4 (1.5) |
| Confidence to quit (0–10) | 6.4 (2.2) | 6.6 (2.1) |
| Strength of tobacco dependency (FTCD; 0–10) | 5.9 (1.8) | 5.9 (1.8) |
| Medium to high (3–6) | 435 (60.4%) | 456 (61.1%) |
| Very high (7–10) | 285 (39.6%) | 290 (38.9%) |

Data are n (%), median (IQR), or mean (SD). SES=Socioeconomic status.
FTCD=Fagerström Test for Cigarette Dependence.

For the primary outcome, self-reported 7-day PPA from combustible tobacco products after 6 months, the rate of smoking cessation was significantly greater in the IG than CG. In the IG, 283 (39.3%) participants reported smoking abstinence while 182 (24.4%) did so in the CG (OR=2.0, 95% CI 1.6–2.5, p<0.0001). This held true for all definitions of smoking, tobacco, or nicotine abstinence (see Table 3). The odds of being abstinent are at least twice as high in the IG than in the CG, independent of the abstinence definition.

**Table 3:** Primary outcome (7-day PPA from combustible tobacco products) and alternative definitions of abstinence as suggested by Piper et al. 2020.(19) Adjusted ORs are from the exploratory logistic regression analyses adjusting for sex, age, SES stratum, Age at which smoking was initiated, FTCD-score, cigarettes per day, desire to quit (baseline), and confidence to quit (baseline); full results of the logistic regression analyses can be found in Table A5 in Appendix F.

|  | IG<br>(n=720)<br>n (%) | CG<br>(n=746)<br>n (%) | OR<br>(95% CI) |  |
| --- | --- | --- | --- | --- |
| 7-day PPA from |  |  | unadjusted | adjusted |
| smoking<br>(combustible tobacco products) | 283<br>(39.3%) | 182<br>(24.4%) | 2.0***<br>(1.6–2.5) | 2.1***<br>(1.7–2.6) |
| tobacco<br>(combustible and smokeless<br>tobacco products, incl. HTP) | 242<br>(33.6%) | 147<br>(19.7%) | 2.1***<br>(1.6–2.6) | 2.1***<br>(1.7–2.7) |
| nicotine<br>(combustible and smokeless<br>tobacco products and ENDS) | 212<br>(29.4%) | 122<br>(16.4%) | 2.1***<br>(1.6–2.8) | 2.2***<br>(1.7–2.9) |

IG=Intervention group; CG=Control group; OR=Odds ratio; PPA=Point-prevalence abstinence; ENDS=Electronic nicotine delivery system; HTP=Heated tobacco products \*\*\*: $p<0.0001$

A similar pattern of results was observed across the secondary outcomes, including the biochemically validated outcome (see Table 4). All 317 (21.6%) participants reporting nicotine abstinence and no use of cannabis products were invited to participate in the cotinine test. Of those, 231 (72.9%) booked a video appointment (IG: 144/198 [72.7%]; CG: 87/119 [73.1%]; p=0.94). In total, 86.1% attended the video appointment (IG: 122/144 [84.7%]; CG: 77/87 [88.5%]; p=0.56). There were 94% (187/199) conclusive tests among those who attended their video appointments. Overall, 53.9% of eligible participants had confirmed abstinence (171/317).

**Table 4:** Secondary outcomes.

| Secondary outcomes | IG<br>n (%) | CG<br>n (%) | p | OR (unadj.)<br>(95% CI) |
| --- | --- | --- | --- | --- |
| Biochemically-validated abstinence | 105 (14.6%) | 66 (8.8%) | <0.0001 | 1.8 (1.3–2.5) |
| Repeated 7-Day PPA | 185 (25.7%) | 104 (13.9%) | <0.0001 | 2.1 (1.6–2.8) |
| Continuous abstinence (allowing max. 5 cigarettes) | 195 (27.1%) | 103 (13.8%) | <0.0001 | 2.3 (1.8–3.1) |

PPA=Point prevalence abstinence; OR=Odds ratio

Among self⍰reported abstainers with conclusive cotinine tests, 171/187 (91.4%) were biochemically verified abstinent (negative predictive value for self⍰reported abstinence), similar across groups (IG: 105/114 [92.1%]; CG: 66/73 [90.4%]; p=0.69). Discordance was 16/187 (8.6%), entirely false negatives (self⍰reported abstinent but cotinine positive). Because biochemical verification was conducted only for self⍰reported abstainers, positive predictive value for self⍰reported smoking and Cohen’s κ are not identifiable without additional verification of self⍰reported smokers.

Of 99 potential serious adverse events (SAEs), contact was achieved for 77. An SAE was confirmed in 62 of these cases (80.5%). The other 22 suspected cases without successful contact were automatically categorised as SAEs. In total, at least one SAE was reported by 31 participants in the IG (4.3%) and 44 participants in the CG (5.9%). No SAE was classified as treatment-related by the trial’s medical lead (CSL). Table A6 in Appendix G gives an overview of SAE categories and their prevalence in each group. As the first sensitivity analysis, the primary endpoint was analysed using multiple imputation with predictive mean matching instead of missing-equals-smoking imputation using all baseline variables as well as intermediary abstinence reports as predictors. This led to a mean estimate of 334.6 (46.5%) in the IG and 214.6 (28.8%) participants in the CG exhibiting smoking abstinence (OR=2.2, 95% CI 1.8–2.8, p<0.0001). In a second set of sensitivity analyses, smoking abstinence rates (again with missing-equals-smoking imputation) were compared within each stratum and sex. This resulted in a statistically significant difference in smoking abstinence rates in favour of the IG within all six subgroups (ORs≥1.6, ps≤0.0154). Because of the low number of users of nicotine replacement therapy or other pharmacological options (IG: 8 [1.1%]; CG: 4 [0.5%]), the effect of pharmacotherapy could not be analysed. A last sensitivity analysis was conducted considering the higher number of active withdrawals in the CG, assuming that all participants actively withdrawing their consent were abstinent. This would result in abstinence rates of 43.2% in the IG and 31.1% in the CG (OR=1.7, 95% CI 1.4–2.1, p<0.0001). Even if assuming this only in the CG and treating active withdrawals in the IG as returning to smoking, the difference in abstinence rates between the groups remained statistically significant (OR=1.4, 95% CI 1.2–1.8, p=0.0010).

Post-hoc analyses of app usage are based on app opening data that were recorded for 711 (98.8%) and data for setting (at least one) quit date that were recorded for 718 (99.7%) participants in the IG. Some data were missing, likely due to connectivity issues or other technical problems. Median app openings were 34 (13–93.5) but participants who reported smoking abstinence at 6 months post-randomisation had substantially more app openings (63 [26–139]) compared to those who did not (22 [8–59]). Most participants (63.5%) set only one quit date, while 14.1% had recorded two quit attempts. Most (89.7%) did not set more than 5 different quit dates.

Similarly, participants in the IG were more satisfied with the app (M=3.7, SD=1.0) than participants in the CG (M=2.4, SD=1.2, t[1221.1]=21.24, p<0.0001, d=1.21, 95% CI 1.09–1.33). Also, the proportion of promoters in the NPS was higher for the IG (30.3%) than for the CG (7.6%), reflecting a significantly higher mean score (6.7 [2.7] vs. 3.6 [3.0], t[1222.6]=19.2, p<0.0001, d=1.09, 95% CI 0.97–1.21).

## Discussion

Smoke Free was found to be effective for smoking cessation for moderately to severely dependent smokers. As a digital intervention, it helped participants achieve and maintain abstinence after 6 months when offered as an adjunct to standard of care in the broad German population and compared to a minimal support control app. The odds of being abstinent were twice as high in the intervention group than in the control group, with self-reported smoking abstinence rates over 39% in the intervention group. Even with a strong performance in the control group, the number needed to treat (NNT) to achieve one additional successful quit using the investigated app was 7, indicating a clinically meaningful effect. This is consistent with previous studies on the potential of digital smoking interventions.(4,6,25) The reported effect size is comparable to conventional face-to-face interventions and cessation services (eg, when they are compared to brief advice and treatment as usual).(26) Results show that digital interventions can be effective. Combined with their inherent scalability and accessibility, they may provide support to patients regardless of temporal or geographical constraints.

This result was yielded from a rigorously designed and conducted RCT with more than 1,450 participants. Like the rigour of the iCanQuit trial of 2,450 participants,(7) the current trial is the largest DiGA trial to date with its double-blind design and high retention rate (>84%). The study complements previous studies of Smoke Free that have focused on the effectiveness of the missions,(12) chatbot,(13) or dragon pet game(14) individually by comparing the full version of the app with versions that lacked the respective feature.

The control app design allowed isolation of the app’s specific efficacy while controlling for technology-related effects, attention, or expectancy bias that could confound results in usual-care-only comparisons. It controls for nonspecific effects such as user engagement, interface usability, and the novelty of app use, isolating the impact of the active therapeutic components from general app-related effects. It also enhances internal validity by reducing confounding variables related to app appearance and functionality, allowing for a more accurate assessment of the intervention’s efficacy. Both groups experienced identical processes of app installation and activation, ensuring comparable technology exposure and minimising differential attrition due to varying digital engagement requirements.

By doing this, this trial directly tackled the widely recognised challenge of suitable control interventions in digital health research, where user expectations and engagement with technology can significantly influence outcomes. We demonstrated that an active control in an RCT in digital smoking cessation is feasible. The comparable attrition rates and robust cessation outcomes in both groups support the rationale for this approach, suggesting such controls are not only attainable but beneficial for the internal and external validity of such a research endeavour. However, the relatively high abstinence rate in the control group suggests that active controls might lead to lower estimates for the treatment effect when comparing it to studies with other forms of control interventions.(4,26) Taken together, these findings may inform future landmark studies and set methodological standards for evaluating digital interventions.

The sample was diverse with respect to age or smoking and cessation history but some limitations remain. First, it comprised more women than men even when smoking (in Germany) is generally more prevalent among men than women.(27) However, epidemiological evidence suggests that women are more likely to attempt to quit and reach out for professional help but less likely to succeed with conventional methods.(28) This means that the sample might be closer to the population of smokers willing to quit and searching for evidence-based methods to do so than smokers in general. Second, relatively few participants were classified as low SES when using the prespecified criterion. Notwithstanding, the underrepresentation of individuals from disadvantaged backgrounds is a common problem in clinical trials.(29) However, Table A3 in Appendix B shows that there is variance in education and income within the sample but parity between both groups, even in these circumstances. Moreover, the difference between both treatment groups is statistically significant even in the relatively small low-SES stratum, making it unlikely that the classification biased the interpretation of results and indicating the effectiveness in this otherwise harder to reach and thus frequently undertreated population group. For future research, there should be an increased focus on recruiting patients from lower SES backgrounds. Third, the number of users of pharmacotherapy is extraordinarily low. This might be because most options are not covered by health insurance and thus pharmacotherapy is relatively unpopular in Germany.(2) It could also be that participants in the trial understood their participation as exclusive and did not use other options concurrently. Fourth and lastly, all participants received the ABC brief intervention and were recruited based on their motivation to quit smoking. This leads to a sample that might be already more invested in their attempt and in the end more successful than when smokers try to quit on their own. While this is representative for the typical DiGA user, it should be considered when comparing trial results internationally and when extrapolating them to the general population.

Self-report measures are prone to bias, but prior evidence indicates that such biases are reduced in remote smoking cessation trials.(30) To further minimise their impact, biochemical validation was employed. Over 91% of the cotinine tests were negative. The primary outcome was also embedded among other smoking-related questions, and participants were repeatedly assured that their responses would not affect further treatment or study participation. Still, because not every eligible participant took part in the cotinine test, this represents a limitation of the study.

Low attrition and the similar retention between groups strengthen the credibility of the findings. While only app openings were available as usage data, they indicated regular use. More granular data (eg, logins per week, time spent with specific features) would have been informative but were not accessible due to the data-minimising requirements of the DiGA framework.

A key strength of the current trial was the inclusion of highly dependent smokers as around 40% exhibited a strong dependence. Furthermore, by restricting the sample to individuals scoring at least 3 points on the FTCD, the sample became more representative of dependent smokers who wish to quit smoking but struggle to do so without any support.

No treatment-related serious adverse events were reported during the study, a finding consistent with previous research on digital behavioural interventions.

Digital interventions hold promise for personalised medicine, particularly through employing just-in-time adaptive interventions (JITAIs).(31) Such interventions adapt to the individual patient’s needs and situation by utilising location- or time-based algorithms (eg, providing tailored feedback based on mood or detecting locations that elicited cravings in the past). Such developments bear the possibility of not only preparing people for cravings but to actively shield people from them.

To quit smoking has a multitude of substantial benefits for the individual(32) and the healthcare system.(33) The Smoke Free app showed to be effective in helping individuals to quit. It doubled the odds of successful abstinence at 6 months after controlling for sociodemographic and smoking-related factors, highlighting its potential as a cost-effective addition to standard care in the German healthcare system.

## Data Availability

Anonymised data, data dictionary, and study material are available online (Research Box 4739; https://researchbox.org/4739&PEER_REVIEW_passcode=JTJBCR).

## Contributors

DC conceived the original idea for the development of the app and provided funding. DC and BPS coordinated the development of the trial apps. PT, TS, SS, and RH wrote the study protocol. AS managed the day-to-day running of the trial, including all participant follow-up. CSL served as the medical lead of the trial. LK and RW undertook and wrote the data analyses. This article was written by LK with input from all coauthors. LK and RW are guarantors for this article. All authors read and approved the final version.

## Transparency declaration

LK and RW (the manuscript’s guarantors) affirm that this manuscript is an honest, accurate, and transparent account of the study being reported; that no important aspects of the study have been omitted; and that any discrepancies from the study as planned (and, if relevant, registered) have been explained.

## Acknowledgments

We thank the tireless contributions of the entire study staff of MEDIACC GmbH. We are very appreciative of the trial participants.

## Summary box

### What is already known on this topic

- Smartphone app-based smoking cessation interventions have shown modest and inconsistent benefits over simpler digital or text message-based comparators in achieving abstinence.
- App-only interventions appear less effective than approaches combining digital interventions with pharmacotherapy.
- Prior trials have been constrained by high attrition rates, reliance on self-reported data, and inconsistent abstinence definitions, leaving uncertainty about app efficacy under rigorous trial conditions.

### What this study adds

- The study suggests that a regulator-approved smoking cessation app can increase abstinence rates compared with usual care.
- The intervention was effective across socioeconomic strata or levels of tobacco dependence and without combination with pharmacotherapy, supporting the use of smoking cessation apps as components of tobacco control strategies.

## References

1. Papadakis S, Katsaounou P, Kyriakos CN, Balmford J, Tzavara C, Girvalaki C, et al. Quitting behaviours and cessation methods used in eight European Countries in 2018: findings from the EUREST-PLUS ITC Europe Surveys. Eur J Public Health. 2020 Jul 1;30(Suppl_3):iii26–33. doi:10.1093/eurpub/ckaa082 PubMed PMID: 32918825; PubMed Central PMCID: PMC7526775.

2. Kotz D, Batra A, Kastaun S. Smoking Cessation Attempts and Common Strategies Employed. Dtsch Arztebl Int. 2020 Jan 6;117(1–2):7–13. doi:10.3238/arztebl.2020.0007 PubMed PMID: 32008606; PubMed Central PMCID: PMC7008148.

3. DEBRA study – Deutsche Befragung zum Rauchverhalten | German Study on Tobacco Use. DEBRA study [Internet]. [cited 2025 Nov 18]. Available from: https://www.debra-study.info/about-debra

4. Guo YQ, Chen Y, Dabbs AD, Wu Y. The Effectiveness of Smartphone App-Based Interventions for Assisting Smoking Cessation: Systematic Review and Meta-analysis. J Med Internet Res. 2023 Apr 20;25:e43242. doi:10.2196/43242 PubMed PMID: 37079352; PubMed Central PMCID: PMC10160935.

5. Naughton F, Nadkarni A. The Contribution of Digital Treatment to Efforts to Reduce Global Tobacco Use. N Engl J Med. 2025 Oct 16;393(15):1449–52. doi:10.1056/NEJMp2500683 PubMed PMID: 41085052.

6. Ubhi HK, Michie S, Kotz D, van Schayck OCP, Selladurai A, West R. Characterising smoking cessation smartphone applications in terms of behaviour change techniques, engagement and ease-of-use features. Transl Behav Med. 2016 Sep;6(3):410–7. doi:10.1007/s13142-015-0352-x PubMed PMID: 27528530; PubMed Central PMCID: PMC4987605.

7. Bricker JB, Watson NL, Mull KE, Sullivan BM, Heffner JL. Efficacy of Smartphone Applications for Smoking Cessation: A Randomized Clinical Trial. JAMA Intern Med. 2020 Nov 1;180(11):1472–80. doi:10.1001/jamainternmed.2020.4055 PubMed PMID: 32955554; PubMed Central PMCID: PMC7506605.

8. Bricker JB, Santiago-Torres M, Mull KE, Sullivan BM, David SP, Schmitz J, et al. Do medications increase the efficacy of digital interventions for smoking cessation? Secondary results from the iCanQuit randomized trial. Addiction. 2024 Apr;119(4):664–76. doi:10.1111/add.16396 PubMed PMID: 38009551; PubMed Central PMCID: PMC10932808.

9. WHO Global Report on Trends in Prevalence of Tobacco Use 2000-2025. 4th ed. Geneva: World Health Organization; 2021. 1 p.

10. Stern AD, Brönneke J, Debatin JF, Hagen J, Matthies H, Patel S, et al. Advancing digital health applications: priorities for innovation in real-world evidence generation. Lancet Digit Health. 2022 Mar;4(3):e200–6. doi:10.1016/S2589-7500(21)00292-2 PubMed PMID: 35216754.

11. Jackson S, Kale D, Beard E, Perski O, West R, Brown J. Effectiveness of the Offer of the Smoke Free Smartphone App Compared With No Intervention for Smoking Cessation: Pragmatic Randomized Controlled Trial. J Med Internet Res. 2024 Nov 15;26:e50963. doi:10.2196/50963 PubMed PMID: 39546331; PubMed Central PMCID: PMC11607577.

12. Crane D, Ubhi HK, Brown J, West R. Relative effectiveness of a full versus reduced version of the ‘Smoke Free’ mobile application for smoking cessation: an exploratory randomised controlled trial. F1000Res. 2018;7:1524. doi:10.12688/f1000research.16148.2 PubMed PMID: 30728950; PubMed Central PMCID: PMC6347038.

13. Perski O, Crane D, Beard E, Brown J. Does the addition of a supportive chatbot promote user engagement with a smoking cessation app? An experimental study. Digit Health. 2019;5:2055207619880676. doi:10.1177/2055207619880676 PubMed PMID: 31620306; PubMed Central PMCID: PMC6775545.

14. White JS, Toussaert S, Raiff BR, Salem MK, Chiang AY, Crane D, et al. Evaluating the Impact of a Game (Inner Dragon) on User Engagement Within a Leading Smartphone App for Smoking Cessation: Randomized Controlled Trial. J Med Internet Res. 2024 Oct 30;26:e57839. doi:10.2196/57839 PubMed PMID: 39475840; PubMed Central PMCID: PMC11561441.

15. Heatherton TF, Kozlowski LT, Frecker RC, Fagerström KO. The Fagerström Test for Nicotine Dependence: a revision of the Fagerström Tolerance Questionnaire. Br J Addict. 1991 Sep;86(9):1119–27. doi:10.1111/j.1360-0443.1991.tb01879.x PubMed PMID: 1932883.

16. Kastaun S, Leve V, Hildebrandt J, Funke C, Klosterhalfen S, Lubisch D, et al. Training general practitioners in the ABC versus 5As method of delivering stop-smoking advice: a pragmatic, two-arm cluster randomised controlled trial. ERJ Open Res. 2021 Jul;7(3):00621–2020. doi:10.1183/23120541.00621-2020 PubMed PMID: 34322552; PubMed Central PMCID: PMC8311138.

17. Cheng CCW, He WJA, Gouda H, Zhang MJ, Luk TT, Wang MP, et al. Effectiveness of Very Brief Advice on Tobacco Cessation: A Systematic Review and Meta-Analysis. J Gen Intern Med. 2024 Jul;39(9):1721–34. doi:10.1007/s11606-024-08786-8 PubMed PMID: 38696026; PubMed Central PMCID: PMC11255176.

18. Bricker JB, Mull KE, McClure JB, Watson NL, Heffner JL. Improving quit rates of web-delivered interventions for smoking cessation: full-scale randomized trial of WebQuit.org versus Smokefree.gov. Addiction. 2018 May;113(5):914–23. doi:10.1111/add.14127 PubMed PMID: 29235186; PubMed Central PMCID: PMC5930021.

19. Piper ME, Bullen C, Krishnan-Sarin S, Rigotti NA, Steinberg ML, Streck JM, et al. Defining and Measuring Abstinence in Clinical Trials of Smoking Cessation Interventions: An Updated Review. Nicotine Tob Res. 2020 Jun 12;22(7):1098–106. doi:10.1093/ntr/ntz110 PubMed PMID: 31271211; PubMed Central PMCID: PMC9633719.

20. West R, Hajek P, Stead L, Stapleton J. Outcome criteria in smoking cessation trials: proposal for a common standard. Addiction. 2005 Mar;100(3):299–303. doi:10.1111/j.1360-0443.2004.00995.x PubMed PMID: 15733243.

21. Adams C, Walpola R, Schembri AM, Harrison R. The ultimate question? Evaluating the use of Net Promoter Score in healthcare: A systematic review. Health Expect. 2022 Oct;25(5):2328–39. doi:10.1111/hex.13577 PubMed PMID: 35985676; PubMed Central PMCID: PMC9615049.

22. JASP - Free and User-Friendly Statistical Software [Internet]. [cited 2025 Nov 18]. JASP - A Fresh Way to Do Statistics. Available from: https://jasp-stats.org/

23. Home Page - Paper Authoring Tool (PAT) [Internet]. [cited 2025 Nov 18]. Available from: https://paperauthoringtool.com/

24. White JS, Salem MK, Toussaert S, Westmaas JL, Raiff BR, Crane D, et al. Developing a Game (Inner Dragon) Within a Leading Smartphone App for Smoking Cessation: Design and Feasibility Evaluation Study. JMIR Serious Games. 2023 Aug 11;11(1):e46602. doi:10.2196/46602

25. Li S, Qu Z, Li Y, Ma X. Efficacy of e-health interventions for smoking cessation management in smokers: a systematic review and meta-analysis. EClinicalMedicine. 2024 Feb;68:102412. doi:10.1016/j.eclinm.2023.102412 PubMed PMID: 38273889; PubMed Central PMCID: PMC10809126.

26. Lancaster T, Stead LF. Individual behavioural counselling for smoking cessation. Cochrane Database Syst Rev. 2017 Mar 31;3(3):CD001292. doi:10.1002/14651858.CD001292.pub3 PubMed PMID: 28361496; PubMed Central PMCID: PMC6464359.

27. Starker A, Kuhnert R, Hoebel J, Richter A. Rauchverhalten und Passivrauchbelastung Erwachsener – Ergebnisse aus GEDA 2019/2020-EHIS. Vol. 7. 2022;7:7–22. doi:10.25646/10290

28. Smith PH, Bessette AJ, Weinberger AH, Sheffer CE, McKee SA. Sex/gender differences in smoking cessation: A review. Prev Med. 2016 Nov;92:135–40. doi:10.1016/j.ypmed.2016.07.013 PubMed PMID: 27471021; PubMed Central PMCID: PMC5085924.

29. Walter JK, Davis MM. Who’s Willing? Characteristics Associated with Willingness to Participate in Clinical Research. IRB: Ethics & Human Res. 2016 Apr 30;38:15–9.

30. Patrick DL, Cheadle A, Thompson DC, Diehr P, Koepsell T, Kinne S. The validity of self-reported smoking: a review and meta-analysis. Am J Public Health. 1994 Jul;84(7):1086–93. doi:10.2105/ajph.84.7.1086 PubMed PMID: 8017530; PubMed Central PMCID: PMC1614767.

31. Naughton F. Delivering ‘Just-In-Time’ Smoking Cessation Support Via Mobile Phones: Current Knowledge and Future Directions. Nicotine Tob Res. 2017 Mar;19(3):379–83. doi:10.1093/ntr/ntw143 PubMed PMID: 27235703.

32. Jackson SE, Jarvis MJ, West R. The price of a cigarette: 20 minutes of life? Addiction. 2025;120(5):810–2. doi:10.1111/add.16757

33. Effertz T. Die Kosten des Rauchens in Deutschland im Jahr 2018 – aktuelle Situation und langfristige Perspektive. Atemwegs-und Lungenkrankheiten. 2019 Jul 1;45(07):307–14. doi:10.5414/ATX02359

