## Appendices for "Effectiveness of the Smoke Free App for Smoking Cessation – Results of RAUCHFREI, a Randomised, Double-blind, Controlled, Two-arm, Parallel, Nationwide, Decentralised, Fully Remote Clinical Trial in Germany"

### Appendix A: Description of the Smoke Free App and the Control App

#### Smoke Free App Features

The Smoke Free DiGA (“Smoke Free – Rauchen aufhören”) consists of multiple interconnected components designed to support smoking cessation through evidence-based behaviour change techniques.

*Onboarding,* the process of introducing new users to an app, takes the form of a *guided setup process* in which new users answer structured questions about their smoking patterns and personal circumstances. Users are asked whether they smoke own-roll tobacco cigarettes or manufactured cigarettes, the cost of their cigarettes (to calculate financial savings), and how soon after waking they first smoke, together with how many cigarettes they smoke per day to enable calculation of the Heaviness of Smoking Index,^1^ a standardised index of cigarette addiction.

The onboarding process also guides users through the process of setting a *personalised quit date*, which can be in the past or in the future based on the user’s readiness to quit and whether they have already begun their quit attempt. Users are asked to *set a savings goal* and to identify their *personal reasons for quitting*, which are stored and can be accessed later to support their motivation to remain abstinent.

The app then creates a *tailored quit plan* based on the individual’s smoking history, dependence level, and quit goals.

Following this, smokers are invited to use the app by following a *structured engagement schedule* based on their quit date.

- In the week before quitting, users are encouraged to engage with preparatory content and set up their personalised plan.
- During the first month post-quit, users receive daily missions: short, evidence-based behavioural tasks assigned each day to help users leave smoking behind, change their identity to that of a non-smoker, feel proud about their achievements, and find new ways of dealing with cravings.
- After the initial 30 days, the missions and notifications are slowly phased out over the next 60 days. Users are prompted via push notifications to engage with the app in the morning with reminder notifications sent around noon and in the evening if users have not interacted with the app.
- Afterwards, their tracking dashboard, health improvements, and savings calculator stay available to users to maintain long-term motivation (see below).

Within this structure, the app features:

- An always-on, comprehensive *tracking dashboard* displaying real-time metrics, including time smoke-free, cigarettes not smoked, money saved, and health improvement indicators across 16 body systems.
- A *craving management system* that includes a diary to record cravings with contextual notes on triggers, mood, and location, pattern analysis to identify high-risk situations, geographic mapping of craving locations, and graphical representations showing craving intensity declining over time.
- A *virtual pet game* to help users maintain their quit by rewarding sustained abstinence as well as providing distraction when cravings arise.
- *Motivational features* such as *unlockable achievement badges* for milestones, options to *share milestones* from within the app, *virtual savings goals* that allow users to designate treats to purchase with money saved, and *personalised motivation reminders* showing user-entered reasons for quitting.
- *Community and social support elements* include an in-app *community chat room* connecting users with peers and access to a *Facebook community grou*p with over 10,000 members, both of which promote peer encouragement.
- An invitation to *log current stress experiences* at random intervals three times a day. In case of spikes in stress experience, *remediating actions within the app* are suggested to prevent stress-induced relapses.
- Check-ins with a *quit coach chatbot*, a rule-based chatbot that implements the *Standard Treatment Programme* from the UK’s National Centre for Smoking Cessation and Training^2^ and is available throughout.

#### Smoke Free App Behaviour Change Techniques

Behaviour Change Techniques (BCTs) are the active ingredients of behaviour change interventions such as the Smoke Free app, specified systematically using agreed terminology, using language that allows for comparison across different interventions, and therefore more accurate evidence synthesis and application. BCTs are the behavioural equivalent of the active ingredients of pharmaceutical formulations and are increasingly recognised as a key part of the specification of behaviour change interventions.^3,4^

The following provides an informal summary of the BCTs and the mechanisms of action that they target, followed by a table listing the BCTs using the formal Behaviour Change Intervention Ontology (BCIO),^4^ which links the BCTs to unique identifiers that can be used for automated search and reasoning, as has become standard in many areas of science and technology.^5^

Relapse to smoking occurs when motivation to smoke in the form of urges or desires (together referred to as cravings) is stronger than the resolve not to smoke in situations in which cigarettes are available or could be acquired.^6^ Therefore, the goal of smoking cessation support is to reduce the incidence and intensity of cravings and achieve a high sustained level of resolve not to smoke, which includes motivation not to smoke, confidence in the ability to sustain abstinence and the self-regulatory capacity to counter cravings. Nicotine withdrawal symptoms, such as mood disturbance, increased appetite, increased anger and difficulty concentrating, contribute to cravings and undermine resolve.^6^ Therefore, the Smoke Free app includes BCTs that aim to target the following mechanisms of action:

- Reduce the incidence of smoking cravings,
- Reduce the intensity of cravings when they occur,
- Reduce the severity and improve coping with nicotine withdrawal symptoms,
- Increase motivation to remain abstinent, and
- Increase the confidence and self-regulatory capacity to maintain abstinence.

The Smoke Free app does this by creatively applying evidence-based principles set out and certified by the UK National Centre for Smoking Cessation and Training (NCSCT) and the German DiGA approval process, requiring demonstration of positive healthcare effects.

*To reduce the incidence of smoking cravings:* Environmental restructuring through missions that prompt users to modify their physical and social environments to reduce smoking cues; trigger identification exercises to recognise and avoid high-risk situations revealed through craving pattern analysis.

*To reduce the intensity of cravings when they occur:* Real-time craving management tools providing immediate distraction (eg, with the virtual pet game) and coping strategies when cravings arise; cognitive reframing techniques delivered through missions and chatbot interactions to change perceptions of cravings; visual feedback through craving graphs showing declining craving intensity over time, which normalises the experience and reinforces that cravings diminish; and a library of evidence-based tips and techniques for resisting specific types of cravings based on context and trigger, following German and international cessation guidelines.

*Reduce the severity of, and improve coping with, nicotine withdrawal symptoms:* Education about expected withdrawal symptoms through the health timeline, which shows when different symptoms typically resolve; behavioural coping strategies taught through missions and check-ins for managing specific withdrawal symptoms such as irritability, restlessness, and difficulty concentrating; and progress monitoring showing health improvements as withdrawal symptoms abate, providing tangible evidence of progress.

*Increase motivation to remain abstinent*: Identity change interventions that help users reframe their self-concept from “smoker” to “non-smoker” through missions; financial savings-tracking with real-time accumulation and virtual treat goals, making abstract benefits concrete and immediately visible throughout the treatment period; health improvement indicators showing 16 different body systems healing on personalised timelines, providing health-based motivation; personal motivation reminders displaying user-entered reasons for quitting at strategic moments; achievement badges and certificates celebrating milestones at intervals consistent with long-term abstinence monitoring; peer support and success stories from the community showing that sustained abstinence is achievable; and motivational messages and quotes delivered through the app to provide ongoing encouragement.

*Increase the confidence and self-regulatory capacity to maintain abstinence*: The “not-a-puff” rule commitment, a behavioural commitment technique reinforcing total abstinence rather than moderation consistent with German and international smoking cessation guidelines; self-monitoring through comprehensive tracking of time spent smoke free, demonstrating accumulated success and building self-efficacy; lapse recovery support providing non-judgmental guidance if users smoke, helping them learn from lapses rather than viewing them as failures; problem-solving training through missions and check-ins that develop specific skills for handling difficult situations; graded tasks starting with manageable challenges and progressively building self-regulatory skills; social support from peers who understand the quit journey and can provide empathetic encouragement and advice; and action planning with personalised strategies developed during check-ins to anticipate and prepare for challenging situations.

Pharmacotherapies have proven effectiveness in aiding smoking cessation.^7^ These include nicotine replacement therapies (NRT), bupropion, and varenicline. They work by reducing the *incidence and intensity of urges to smoke and the severity of withdrawal symptoms*. Therefore, the Smoke Free app provides information about pharmacological support, including educational content about NRT products, including transdermal patches, nicotine chewing gum, lozenges, inhalers, and nasal sprays. It includes guidance on combining NRT products for maximum effectiveness, consistent with German cessation guidelines. It includes information about varenicline and bupropion. It also includes support for medication adherence through behavioural techniques integrated into the app, as well as coordination with the German healthcare system to facilitate obtaining necessary prescriptions and obtaining reimbursement.

#### Formal BCT Coding of the Smoke Free App using the Behaviour Change Technique Ontology

Below is a list of unique Behaviour Change Techniques delivered by the Smoke Free app specified using the Behaviour Change Technique Ontology (BCTO). The mapping of app content to BCTs was undertaken by LK and checked and revised by RW.

| **Broad category of BCT** | **BCT Name** | **BCT ID** |
| --- | --- | --- |
| Goal setting | set measurable behaviour goal | BCIO:007300 |
|  | set measurable outcome goal | BCIO:007301 |
|  | affirm commitment | BCIO:007015 |
|  | make a goal public | BCIO:007122 |
|  | action planning | BCIO:007010 |
|  | goal strategising | BCIO:007008 |
|  | attend to discrepancy between current behaviour and goal | BCIO:007012 |
|  | review behaviour goal | BCIO:007011 |
|  | review behaviour goal plan | BCIO:007299 |
| Goal reminding | advise to keep behaviour goal in mind | BCIO:007141 |
|  | advise to keep outcome goal in mind | BCIO:007142 |
| Advising on behaviour | context-specific non-enactment of behaviour | BCIO:007169 |
|  | context-specific repetition of alternative behaviour | BCIO:007097 |
|  | facilitate alternative goal-directed activity | BCIO:007171 |
|  | increase awareness of option of novel behaviour | BCIO:007174 |
|  | set graded tasks | BCIO:007100 |
| Advising on environmental restructuring | advise to avoid people who do an unwanted behaviour | BCIO:050330 |
|  | reduce exposure to cues for the behaviour | BCIO:007153 |
|  | remove aversive stimulus | BCIO:050331 |
| Advising on how to do a behaviour | guide how to perform behaviour | BCIO:007050 |
|  | suggest how to perform behaviour | BCIO:007303 |
| Advising on social support | advise to seek appraisal support | BCIO:007033 |
|  | advise to seek emotional support | BCIO:007031 |
|  | advise to seek informational support | BCIO:007032 |
|  | advise to seek instrumental support | BCIO:007030 |
| Arranging social support | arrange appraisal support | BCIO:007038 |
|  | arrange emotional support | BCIO:007036 |
|  | arrange informational support | BCIO:007037 |
|  | arrange instrumental support | BCIO:007035 |
| Confidence building | persuade about personal capability | BCIO:007137 |
|  | prompt focus on past success | BCIO:007139 |
|  | prompt self-talk | BCIO:007140 |
|  | prompt thinking related to successful performance | BCIO:007239 |
|  | remind about personal capability | BCIO:007060 |
| Focusing on self-identity | adopt changed self-identity | BCIO:007160 |
|  | adopt positive self-identity | BCIO:007160 |
|  | affirm valued self-identity | BCIO:007159 |
| Improving credibility | present information from credible influence | BCIO:007075 |
| Informing about consequences | inform about negative emotional consequences | BCIO:007177 |
|  | inform about positive emotional consequences | BCIO:007181 |
|  | inform about positive health consequences | BCIO:007183 |
| Making consequences salient | increase salience of consequences | BCIO:007068 |
|  | induce anticipated regret | BCIO:007067 |
|  | prompt comparative imagining of future outcomes | BCIO:007070 |
| Managing mental processes | advise behavioural ways to reduce negative emotions | BCIO:050333 |
|  | advise cognitive ways to increase positive emotions | BCIO:050334 |
|  | advise cognitive ways to reduce negative emotions | BCIO:050337 |
|  | advise distraction | BCIO:007154 |
|  | conserve mental resources | BCIO:007134 |
|  | enable person to manage automatic responses | BCIO:007143 |
|  | provide distraction | BCIO:007155 |
| Monitoring and feedback | monitor emotional consequences | BCIO:007066 |
|  | provide feedback on behaviour | BCIO:007023 |
|  | provide feedback on outcome of behaviour | BCIO:007027 |
|  | self-monitor behaviour | BCIO:007024 |
|  | self-monitor outcome of behaviour | BCIO:007025 |
| Perspective changing | draw attention to incompatible beliefs | BCIO:007057 |
|  | inform about antecedents | BCIO:007052 |
|  | re-attribute cause | BCIO:007053 |
|  | reframe past behaviour | BCIO:007056 |
|  | suggest different perspective on behaviour | BCIO:007302 |
| Promoting pharmacological support | encourage pharmacological support | BCIO:007146 |
| Rewarding | plan inclusion of enjoyment | BCIO:007061 |
|  | promise positive consequence for alternative behaviour | BCIO:007251 |
|  | promise positive consequence for behaviour | BCIO:007202 |
|  | promise positive consequence for outcome of behaviour | BCIO:007216 |
|  | provide positive social consequence for alternative behaviour | BCIO:007266 |
|  | provide positive social consequence for behaviour | BCIO:007265 |
|  | provide positive social consequence for outcome of behaviour | BCIO:007271 |
| Using associative learning | associative learning to elicit behaviour | BCIO:007091 |
|  | associative learning to extinguish behaviour | BCIO:007092 |

Table A1: BCT Coding of the Smoke Free DiGA.

#### Description of the Control App

Participants in the control group received access to a reduced, static version of the app for 6 months that was identical in design on a smartphone’s home screen:

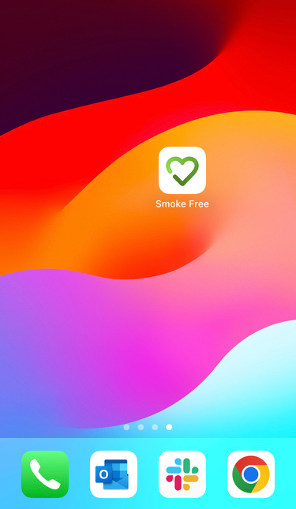

It mirrored the design of the app in the intervention group but only offered static, text-based smoking cessation information modelled on flyers common in GP offices. It provided information useful in aiding the participants’ quit attempts under four headings.

##### Health

##### Restore your health

- As soon as you put out your last cigarette, your body begins to repair the damage.
- Your blood pressure improves, your pulse slows down, and the risk of a heart attack decreases. It won't be long before you notice that your sense of taste and smell are also improving.
- When you breathe in, your improved circulation will oxygenate your whole body, so you'll have a lot more energy as well.

##### Cravings

##### Deal with the craving

Learn to resist cravings and you increase your chances of success. Snacks reduce the intensity and frequency of cravings. Along with the techniques we are going to teach you, you will be well equipped to fight every single craving that arises. But did you know that certain foods can also help?

- Yoghurt (contains calcium and protein) with fruit is a sweet treat.
- Hummus is a food that is high in nutrients, fibre, and protein. Paired with crunchy veggies like carrots and celery sticks, it's a snack that's (almost) a meal in itself.
- Try frozen grapes. They reduce cravings for sweets, and because they're frozen, they take longer to eat.
- Popcorn is a high-fiber whole grain product that is relatively low in calories. It's a great snack that satisfies hunger and keeps your hands busy.

##### Exercise Tab

##### The power of movement

- Exercise quells hunger pangs and keeps you from gaining weight, something many dread most. It's also a great way to improve your mood and manage stress.
- You don't have to run a marathon, you don't even have to go to the gym. Any kind of physical activity relieves tension and reduces the urge to smoke.
- Jump a little, walk up and down the stairs, touch your toes ten times, or go for a walk in the fresh air. Be proud of how much good you are doing for your body.
- Some people (because of a disability or illness) cannot practise every sport. But there are still many activities you can enjoy to take your mind off smoking and get fitter.
- Once you've found something you enjoy, you should try a little harder each day to achieve new goals. Be careful not to exceed your limits! If you are unsure, you can always consider seeking professional advice.

##### Tips

##### Our top tips

- Water is surprisingly good at suppressing cravings. Drink lots of it.
- A glass of milk makes cigarettes taste bad and is also high in nutrients. A warm glass of milk can help you fall asleep.
- Eat more fruits, vegetables, and whole grains. People who smoke tend to eat less than non-smokers. So make sure you eat a balanced diet.
- Cut down on caffeine and alcohol. While they give you a brief boost of energy, if you're not getting enough nutrients, you'll eventually end up feeling drained.
- The body needs a lot of energy to digest a large meal. You may feel tired afterwards, which may make it hard to resist the craving. Instead, eat several smaller meals throughout the day.
- Snacking on unsalted nuts and seeds helps reduce fatigue and hunger, as well as the craving for a cigarette.

#### Formal BCT Coding of the Control App using the Behaviour Change Technique Ontology

Below is a list of unique Behaviour Change Techniques delivered by the control app specified using the Behaviour Change Technique Ontology (BCTO). The mapping of app content to BCTs was undertaken by LK and checked and revised by RW.

| **Broad category of BCT** | **BCT Name** | **BCT ID** |
| --- | --- | --- |
| Advising on behaviour | suggest to change behaviour | BCIO:007076 |
|  | advise specific behaviour | BCIO:007168 |
| Informing about consequences | increase awareness of consequences | BCIO:007062 |
| Improving credibility | present information from credible influence | BCIO:007075 |
| Focusing on self-identity | prompt focus on self-identity | BCIO:007157 |
| Advising on how to do a behaviour | guide how to perform behaviour | BCIO:007050 |
| Advising on environmental restructuring | restructure the environment | BCIO:007150 |
| Confidence building | prompt focus on past success | BCIO:007139 |

Table A2: BCT Coding of the Control App.

#### References for Appendix A

1	Heatherton TF, Kozlowski LT, Frecker RC, Rickert W, Robinson J. Measuring the Heaviness of Smoking: using self-reported time to the first cigarette of the day and number of cigarettes smoked per day. *British Journal of Addiction* 1989; **84**: 791–800.

2	Standard Treatment Programme. https://www.ncsct.co.uk/publications/ncsct-standard-treatment-programme (accessed Nov 17, 2025).

3	Accelerating Social and Behavioral Science Through Ontology Development and Use | National Academies. https://www.nationalacademies.org/our-work/accelerating-social-and-behavioral-science-through-ontology-development-and-use (accessed Nov 17, 2025).

4	Marques M, Wright A, Corker E, *et al.* The Behaviour Change Technique Ontology: Transforming the Behaviour Change Technique Taxonomy v1 [version 2; peer review: 4 approved]. *Wellcome Open Research* 2024; **8**.

5	Uschold M, Gruninger M. Ontologies: principles, methods and applications. *The Knowledge Engineering Review* 1996; **11**: 93–136.

6	West R. The Multiple Facets of Cigarette Addiction and What They Mean for Encouraging and Helping Smokers to Stop. *COPD: Journal of Chronic Obstructive Pulmonary Disease* 2009; **6**: 277–83.

7	Lindson N, Theodoulou A, Ordóñez-Mena JM, *et al.* Pharmacological and electronic cigarette interventions for smoking cessation in adults: component network meta-analyses. *Cochrane Database Syst Rev* 2023; **9**: CD015226.

### Appendix B: Composite measure of socioeconomic status

|  | **Score** | **IG (n=720)** | **CG (n=746)** |
| --- | --- | --- | --- |
| Education |  |  |  |
| no school-leaving certificate | 1 | 2 (0.3%) | 7 (0.9%) |
| basic secondary school leaving certificate | 2 | 48 (6.7%) | 34 (4.6%) |
| intermediate secondary school leaving certificate | 3 | 209 (29.0%) | 218 (29.2%) |
| entrance qualification for universities of applied sciences | 4 | 141 (19.6%) | 143 (19.2%) |
| general university entrance qualification | 5 | 320 (44.4%) | 344 (46.1%) |
| Household income per capita [€] |  |  |  |
| 0–999 | 0.5 | 75 (10.4%) | 73 (9.8%) |
| 1,000–1,999 | 1.0 | 183 (25.4%) | 193 (25.9%) |
| 2,000–2,999 | 1.5 | 252 (35.0%) | 242 (32.4%) |
| 3,000–3,999 | 2.0 | 113 (15.7%) | 133 (17.8%) |
| 4,000–4,999 | 2.5 | 46 (6.4%) | 51 (6.8%) |
| 5,000–5,999 | 3.0 | 25 (3.5%) | 28 (3.8%) |
| 6,000–6,999 | 3.5 | 10 (1.4%) | 9 (1.2%) |
| 7,000–7,999 | 4.0 | 2 (0.3%) | 7 (0.9%) |
| 8,000–8,999 | 4.5 | 5 (0.7%) | 3 (0.4%) |
| 9,000–9,999 | 5.0 | 0 (0%) | 1 (0.1%) |
| 10,000–10,999 | 5.5 | 1 (0.1%) | 1 (0.1%) |
| ≥11,000 | 6.0 | 8 (1.1%) | 5 (0.7%) |

Data are scores or n (n, CG). A sum score of education and household income per capita was used to classify participants into low (≤3.5) or medium-high (>3.5) socioeconomic status.

Table A3: Socioeconomic status classification.

### Appendix C: Assessments at each visit

| **Measure** | **Screening** | **Baseline (Day 0)** | **FU1 (Day 30)** | **FU2 (Day 90)** | **FU3 (Day 180)** |
| --- | --- | --- | --- | --- | --- |
| Inclusion/exclusion criteria incl. age, FTCD | x |  |  |  |  |
| Comorbidities, medication, and potential (S)AEs | x | x | x | x | x |
| Demographics: sex, relationship status, education, household income |  | x |  |  |  |
| Smoking-related background: age at which smoking was initiated, years of smoking, cigarettes smoked per day, number of quit attempts |  | x |  |  |  |
| Smoking household |  | x |  |  | x |
| 7-Day PPAs of a) combustible tobacco products, b) electronic nicotine delivery systems, c) heated tobacco products, d) non-combustible tobacco products |  | x | x | x | x |
| 30-Day PPA any smoking product; smoked anything since quit date |  |  | x | x | x |
| Experience of withdrawal symptoms in past attempts/during the trial; use of support (medication, apps, behavioural therapy) in past attempts/during the trial |  | x | x | x | x |
| Desire to quit, confidence to quit |  | x | x | x | x |
| NPS, app satisfaction, usage data (automatically retrieved) |  |  |  |  | x |
| Other exploratory outcomes (general well-being, stress experience, anxiety/depression, use of cannabis products) |  | x |  |  | x |
| 30-Day PPA psychotropic drugs, narcotics, or sedatives |  | x | x | x | x |

Table A4: Overview of follow-up intervals and assessments done at each visit.

### Appendix D: Cotinine test procedure

Biochemical verification was offered only to participants who self-reported 7-day abstinence from all tobacco products and electronic nicotine delivery systems (ENDS) and cannabis products at the last follow-up (FU3). Participants were informed about the procedure and the additional €30 compensation. Because FU3 assessed use of nicotine replacement therapy (NRT) only with reference to use alongside the study participation but not limited to the last 7 days, it was not used to preclude participation in the cotinine test.

Participants who agreed booked a video consultation at a convenient time. The study team sent out the cotinine test kit at the earliest opportunity. The median time from booking to sending out the test was 1 day (0–3). The median time from sending out to testing was 5 days (5–7), yielding a median interval of 6 days (5–9) between FU3 completion and cotinine testing.

All tests used the same NarcoCheck^®^ Saliva Screening Test (Kappa City Biotech SAS, 32 rue Danton, 03100 Montluçon, France). It is an immuno-chromatographic test designed to detect nicotine intake until up to 72 hours after the last intake and has a cut-off of 20 ng/mL. Its result is qualitative and according to the manufacturer, it returns a positive result when nicotine replacement products are used but not when one is exposed to second-hand smoke. Thus, a positive result in this test indicates active nicotine use. This is in line with saliva cotinine levels reported in the literature.^1^ For this device, the manufacturer does not report sensitivity or specificity in the instructions for use but representative saliva tests using cut-offs of 10–20 ng/mL typically show high sensitivity and specificity versus reference methods.^2^

In 50.0% of the positive test results, participants reported recent NRT or ENDS use as the reason for a positive test result. However, per the statistical analysis plan, any positive cotinine test result was classified as non-abstinent for the secondary outcome measure independent of self-reported NRT/ENDS use. The same was true for any test failing to produce any or a valid test result. This means that only negative test results were classified as abstinent.

#### References for Appendix D

1	Etzel RA. A review of the use of saliva cotinine as a marker of tobacco smoke exposure. *Prev Med* 1990; **19**: 190–197.

2	Kim S. Overview of Cotinine Cutoff Values for Smoking Status Classification. *Int J Environ Res Public Health* 2016; **13**: 1236.

### Appendix E: Power calculation

A systematic literature search during the study’s conception identified three RCTs of mobile interventions comparable to the Smoke Free app in terms of functionality and complexity of the therapeutic approach. All three were conducted in the US;^1–3^ there was no comparable RCT conducted in Germany at the time. As a conservative estimate for the intervention group, we took the lowest out of the abstinence rates observed in the intervention group, namely Craving to Quit’s 18%.^3^

For the control group, we looked at abstinence rates observed in studies administering ABC brief advice in the German healthcare context (11%)^4^ and comparable control groups in a Cochrane Review of other mobile app trials.^5^ We identified a quit rate of 12% for a comparable minimal support intervention.^6^ We took the higher value of 12% as our estimate for the abstinence rate of the control group.

#### References for Appendix E

1	Bricker JB, Watson NL, Mull KE, Sullivan BM, Heffner JL. Efficacy of smartphone applications for smoking cessation: A randomized clinical trial. *JAMA Intern Med* 2020; **180**: 1472–1480.

2	Danaher BG, Tyler MS, Crowley RC, Brendryen H, Seeley JR. Outcomes and Device Usage for Fully Automated Internet Interventions Designed for a Smartphone or Personal Computer: The MobileQuit Smoking Cessation Randomized Controlled Trial. *JMIR* 2019; **21**: e13290.

3	Garrison KA, Pal P, O'Malley SS, et al. Craving to Quit: A Randomized Controlled Trial of Smartphone App-Based Mindfulness Training for Smoking Cessation. *Nicotine Tob Res* 2020; **22**: 324–331.

4	Kastaun S, Viechtbauer W, Leve V, et al. Quit attempts and tobacco abstinence in primary care patients: follow-up of a pragmatic, two-arm cluster randomised controlled trial on brief stop-smoking advice – ABC versus 5As. *ERJ Open Res* 2021; **180**: 00224-2021.

5	Whittaker R, McRobbie H, Bullen C, Rodgers A, Gu Y, Dobson R. Mobile phone text messaging and app-based interventions for smoking cessation. *Cochrane Database Syst Rev* 2019; **10**: CD006611.

6	Cobos-Campos R, Apiñaniz Fernández de Larrinoa A, Sáez de Lafuente Moriñigo A, Parraza Diez N, Aizpuru Barandiaran F. Effectiveness of Text Messaging as an Adjuvant to Health Advice in Smoking Cessation Programs in Primary Care. A Randomized Clinical Trial. *Nicotine Tob Res* 2017; **19**: 901–907.

### Appendix F: Results of the logistic regressions for smoking abstinence (primary outcome) and tobacco and nicotine abstinence (exploratory outcomes)

|  |  | **smoking abstinence** | | **tobacco abstinence** | | **nicotine abstinence** | |
| --- | --- | --- | --- | --- | --- | --- | --- |
| **Model** | **Variable** | **OR (95% CI)** | **p** | **OR (95% CI)** | **p** | **OR (95% CI)** | **p** |
| 0 | (Intercept) | 0.32 (0.27–0.38) | <0.0001 | 0.25 (0.20–0.29) | <0.0001 | 0.20 (0.16–0.24) | <0.0001 |
|  | Intervention (unadjusted) | 2.01 (1.60–2.51) | <0.0001 | 2.06 (1.63–2.62) | <0.0001 | 2.13 (1.66–2.75) | <0.0001 |
| A | (Intercept) | 0.17 (0.07–0.44) | 0.0002 | 0.08 (0.03–0.23) | <0.0001 | 0.06 (0.02–0.17) | <0.0001 |
|  | Sex (female) | 0.97 (0.77–1.22) | 0.7746 | 0.94 (0.74–1.20) | 0.6392 | 1.00 (0.77–1.29) | 0.9935 |
|  | Age | 1.00 (0.99–1.01) | 0.5808 | 1.00 (0.98–1.01) | 0.4172 | 1.00 (0.99–1.01) | 0.9792 |
|  | SES (high) | 1.33 (0.86–2.07) | 0.1980 | 1.32 (0.82–2.10) | 0.2527 | 1.33 (0.81–2.20) | 0.2622 |
|  | Age at which smoking was initiated | 1.01 (1.00–1.02) | 0.2984 | 1.01 (1.00–1.02) | 0.1787 | 1.00 (0.99–1.02) | 0.4343 |
|  | Cigarettes smoked per day | 1.00 (0.98–1.02) | 0.7793 | 1.01 (0.98–1.03) | 0.5635 | 1.01 (0.98–1.03) | 0.6199 |
|  | Desire to quit | 1.11 (1.02–1.21) | 0.0134 | 1.15 (1.05–1.25) | 0.0034 | 1.15 (1.04–1.26) | 0.0055 |
|  | Confidence to quit | 1.08 (1.02–1.14) | 0.0098 | 1.11 (1.05–1.18) | 0.0005 | 1.13 (1.06–1.20) | 0.0003 |
|  | Strength of tobacco dependency (FTCD) | 0.89 (0.83–0.96) | 0.0035 | 0.88 (0.82–0.96) | 0.0030 | 0.87 (0.80–0.95) | 0.0015 |
| B | (Intercept) | 0.10 (0.04–0.27) | <0.0001 | 0.05 (0.02–0.14) | <0.0001 | 0.03 (0.01–0.10) | <0.0001 |
|  | Sex (female) | 0.98 (0.77–1.23) | 0.8373 | 1.00 (0.98–1.01) | 0.4382 | 1.00 (0.99–1.01) | 0.9872 |
|  | Age | 1.00 (0.99–1.01) | 0.6069 | 1.01 (1.00–1.02) | 0.1893 | 1.00 (0.99–1.02) | 0.4633 |
|  | SES (high) | 1.42 (0.91–2.21) | 0.1266 | 1.01 (0.98–1.03) | 0.6489 | 1.00 (0.98–1.03) | 0.7101 |
|  | Age at which smoking was initiated | 1.01 (0.99–1.02) | 0.3076 | 1.14 (1.04–1.25) | 0.0053 | 1.14 (1.03–1.26) | 0.0085 |
|  | Cigarettes smoked per day | 1.00 (0.98–1.02) | 0.8869 | 1.13 (1.06–1.20) | 0.0001 | 1.14 (1.07–1.22) | <0.0001 |
|  | Desire to quit | 1.11 (1.02–1.20) | 0.0197 | 0.89 (0.82–0.97) | 0.0056 | 0.88 (0.80–0.96) | 0.0030 |
|  | Confidence to quit | 1.09 (1.03–1.16) | 0.0034 | 0.95 (0.74–1.22) | 0.7020 | 1.01 (0.78–1.31) | 0.9283 |
|  | Strength of tobacco dependency (FTCD) | 0.90 (0.83–0.97) | 0.0063 | 1.41 (0.87–2.27) | 0.1612 | 1.43 (0.86–2.39) | 0.1652 |
|  | Intervention (adjusted) | 2.07 (1.65–2.60) | <0.0001 | 2.15 (1.68–2.74) | <0.0001 | 2.23 (1.72–2.88) | <0.0001 |

Table A5: Results from the logistic regression for the primary outcome *smoking abstinence* (self-reported 7-day PPA from combustible tobacco products) and the exploratory outcomes *tobacco abstinence* (self-reported 7-day PPA from smokeless and combustible tobacco products) and *nicotine abstinence* (self-reported 7-day PPA from all tobacco products and electronic nicotine delivery systems).

### Appendix G: Serious Adverse Events

| **SAE category** | **total** | **possibly related** |
| --- | --- | --- |
| Mental illnesses | 20 (13) | 20 (13) |
| Affective disorders (occupational burnout, depression) | 10 (6) | 10 (6) |
| Panic disorder or panic attacks | 4 (2) | 4 (2) |
| Schizophrenia/ schizoaffective disorder | 3 (2) | 3 (2) |
| Personality disorders | 1 (1) | 1 (1) |
| Posttraumatic stress disorder | 1 (0) | 1 (0) |
| Stress-related increase in blood pressure | 1 (1) | 1 (1) |
| Without further information | 3 (2) | 3 (2) |
| Physical illnesses | 60 (32) | 1 (1) |
| Emergency operation (eg, appendicitis, cyst removal, kidney stones) | 9 (5) | - |
| Operation (without further information) | 8 (4) |  |
| Lung disease (eg, asthma, COPD, pneumonia) | 4 (2) |  |
| Planned operation (eg, gastric band adjustment, hip replacement) | 4 (2) |  |
| Tissue hernias (abdominal wall, groin, navel, diaphragm) | 4 (0) |  |
| New diagnosis of a chronic disease (eg, epilepsy, multiple sclerosis) | 4 (2) |  |
| Circulatory disorder (aneurysm, PAD, thrombosis) | 3 (1) |  |
| Cardiovascular disease (eg, heart attack, cardiac arrest) | 3 (1) |  |
| Bone fractures | 3 (2) |  |
| Cancer diseases | 3 (1) |  |
| Accidents | 3 (3) |  |
| Acute inflammation (renal pelvis) or sepsis | 2 (1) |  |
| Intervertebral disc disease | 2 (2) |  |
| Diabetes | 2 (2) |  |
| Planned operation with longer follow-up care | 2 (1) |  |
| Fibromyalgia | 1 (1) |  |
| Colic | 1 (0) |  |
| Long COVID or ME/CFS | 1 (1) | 1 (1) |
| Sleep apnea | 1 (1) |  |
| Pregnancy-related events | 4 (3) |  |
| Miscarriage | 3 (2) |  |
| Ectopic pregnancy | 1 (1) |  |

Data are N (n, CG). COPD=Chronic obstructive pulmonary disease; PAD=Peripheral artery disease; Long COVID=Group of health problems persisting after an initial period of Coronavirus disease 2019 infection; ME/CFS= Myalgic encephalomyelitis/chronic fatigue syndrome.

Table A6: SAE classification.
